# COVID-19 detection on IBM quantum computer with classical-quantum transfer learning

**DOI:** 10.1101/2020.11.07.20227306

**Authors:** Erdi Acar, İhsan Yilmaz

## Abstract

Diagnose the infected patient as soon as possible in the coronavirus 2019 (COVID-19) outbreak which is declared as a pandemic by the world health organization (WHO) is extremely important. Experts recommend CT imaging as a diagnostic tool because of the weak points of the nucleic acid amplification test (NAAT). In this study, the detection of COVID-19 from CT images, which give the most accurate response in a short time, was investigated in the classical computer and firstly in quantum computers. Using the quantum transfer learning method, we experimentally perform COVID-19 detection in different quantum real processors (IBMQx2, IBMQ-London and IBMQ-Rome) of IBM, as well as in different simulators (Pennylane, Qiskit-Aer and Cirq). By using a small number of data sets such as 126 COVID-19 and 100 Normal CT images, we obtained a positive or negative classification of COVID-19 with 90% success in classical computers, while we achieved a high success rate of 94-100% in quantum computers. Also, according to the results obtained, machine learning process in classical computers requiring more processors and time than quantum computers can be realized in a very short time with a very small quantum processor such as 4 qubits in quantum computers. If the size of the data set is small; Due to the superior properties of quantum, it is seen that according to the classification of COVID-19 and Normal, in terms of machine learning, quantum computers seem to outperform traditional computers.

## 1. Introduction

The coronavirus outbreak (COVID-19), which occurred in China in December 2019, spread rapidly around the world and was declared a pandemic by the world health organization (WHO)[1]. It is extremely important to be able to diagnose COVID-19 in an infected patient during the pandemic process.

In order to diagnose COVID-19, it is expected that the nucleic acid amplification test (NAAT) of the respiratory tract or blood samples will yield positive results using reverse transcription real-time fluorescence polymerase chain reaction (RT-PCR)[2]. However, as a result of the current clinical experiences, the detection rate and sensitivity are low due to the low viral load in the early stage. Therefore, it is inevitable to give wrong results. Besides, it can give only positive or negative results. The severity and progression of the infection cannot be monitored. It may take 1 day or more to determine the test result after taking the test sample from the patient. In a patient with suspected COVID-19, the test results are negative, but if findings are found in the CT imaging results, the patient should be quarantined and treated as soon as possible[3]. This is very important to help decision making of clinicians for quick isolation and appropriate patient treatment since computerized tomography (CT) which gives both fast and accurate results has a prognostic role in the early diagnosis of COVID-19[4]. The advantage of CT imaging is obvious in the diagnosis of COVID-19. For these reasons, experts recommend CT imaging as the main basis for the diagnosis of COVID-19[5]. Because of a large number of infected patients and the heavy workload on healthcare workers, the computer-aided with machine learning algorithms system can accelerate the diagnosis process.

Deep learning has widespread applications in solving real-world problems such as autonomous vehicles, natural language processing, computer vision, biological analysis, and so on. A large amount of data is required for the efficient operation of deep learning. On the other hand, access to medical data often requires special permission, so access to medical data is not always possible. Therefore, the size of medical data sets is small. When the size of the data set is small, data can be augmentation with generative adversarial network, or transfer learning is used[6–9]. In addition to such a case, quantum machine learning can make the machine learning performance increase and efficient due to the superior properties of quantum such as entanglement and superposition [10, 11]. The superposition is evaluation of all possible states of a qubit state at the same time. If the two-qubit states are interrelated and this relationship is valid even at infinite distance, the two-qubit states are entangled with each other. This concept can be explained as the effect of any operation performed in one qubit on the other qubit instantly[12, 13].

Both machine learning and quantum computing are expected to play a role in how society processes information. These two areas are explored in the quantum machine learning discipline. Quantum machine learning progresses with opportunities in the development of quantum computers. In the context of quantum machine learning, related researches are conducted on whether quantum information brings a new perspective on how machines recognize patterns in the data, whether they can learn from fewer training data, their ability to develop new machine learning methods, or to determine the components and bottlenecks of quantum machine learning algorithms[11, 14]. There are two different strategies when designing quantum machine algorithms. The first is to translate classical machine learning models into quantum computing language to achieve an algorithmic acceleration. The second is to combine quantum tools that look like the results of classical algorithms, keeping computing resources low[11, 14, 15].

Quantum machine learning can be used for machine learning by a completely new type of computing device, a quantum computer. Quantum machine learning is predicted to be superior in performance compared to conventional machine learning if data are obtained as quantum and noise rates in quantum processors are minimized[16]. Some research focuses on ideal, universal quantum computers. However, there has been a rapidly growing interest in quantum machine learning in near-term quantum devices.

There are many studies in the literature on quantum machine learning. Quantum computers produce non-typical model patterns that are thought to not produce conventional computers efficiently. For this reason, it is foreseen that quantum computers can perform more effectively than classical computers in terms of machine learning. In this context, machine learning is studied with a large number of vectors in the high dimensional property space and there is a lot of computational complexity. In order to solve this problem, Lloyd et al.[17], a study has been made on supervised and unsupervised quantum machine learning that there may be an exponential acceleration in both the number and size of vectors. The quantum support vector machine algorithm has been developed by Rebentrost et al.[18] for binary classification and they have achieved a logarithmic acceleration compared to the classical support vector machine. This study also plays an important role in data privacy. How to design and apply quantum machine learning algorithms working on quantum computers was discussed by Biamonte et al.[14]. Giovannetti et al.[19] pioneered how to represent classical data in the language of quantum mechanics through the study of quantum random access memory (QRAM). Quantum nearest neighbor algorithm has been developed for classification by Wiebe et al.[20]. Grant et al.[21] performed binary classification tasks coded into quantum states using the popular Iris and MNIST dataset with hierarchical quantum circuits on the ibmqx4 quantum computer. Kapoor et al.[22] suggested quantum perceptron models compared to the classic perceptron and showed that it would provide demonstrable accelerations. Farhi and Neven [23] study classification with quantum neural networks on near term processors. Cai et al.[24], experimentally demonstrated the suitability of quantum machine learning within the framework of quantum controlled and unsupervised learning, by using a photonic quantum computer, manipulating high dimensional vectors on a double basis, and the distance between the vectors can be estimated.

In the hybrid classical-quantum machine learning, where the data are classic, the data are first coded into the quantum language and the operations are done with quantum operators. In this context, Mari et al.[25] investigated transfer learning in hybrid classical-quantum neural networks. Zen et al.[26] used transfer learning for the scalability of neural network quantum states. Piat et al.[27] studied image classification with quantum pre-training and auto-encoders. Verdon et al.[28] examined learning to learn with quantum neural networks via classical neural networks. Henderson et al.[29] proposed hybrid quantum convolution neural network architecture.

The main purpose and contributions of this study: (1) From Computerized Tomography (CT) images, we propose to diagnose COVID-19 infected patients and Normal(healthy) patients with a hybrid quantum machine learning algorithm. (2) Using the quantum transfer learning method, we experimentally perform COVID-19 detection on different quantum real processors (IBMQx2, IBMQ-London and IBMQ-Rome) and on different noisy simulators (Pennylane, Qiskit-Aer and Cirq). (3) We perform the machine learning process, which requires more processors and time in classical computers, by training a variational quantum circuit with a very small quantum processor such as 4 qubits. (4) In this study, since the CT images are classic, we perform image processing and feature extraction operations on the data with classical methods. (5) We propose an effective circuit diagram consisting of quantum gates to encode classical data into the quantum circuit. (6) When the data set is small, we compare the performance results of the classical and quantum model in terms of accuracy, precision, recall, f1-score and specificity. (7) We show that quantum computers are advantageous in terms of classification of COVID-19 and Normal CT images in the case of a small dataset due to the superior properties of quantum (superposition and entanglement). (8) We think that this study is a pioneer in the realization of COVID-19 detection using quantum machine learning.

The rest of the paper is organized as follows. In Section 2, the materials and methods used in the study are presented. Section 3 presents the results of classic and hybrid models according to different performance metrics. In Section 5, the results obtained from the study were discussed.

## 2. Matarial and Method

### 2.1. Dataset

The data set we used in our study was obtained from open source databases and combining the datasets. ^1,2,3,4,5,6,7^. The obtained data was weed up and a clean dataset was created. The resulting dataset consists of a total of 2658 lung CT images, 1296 COVID-19 and 1362 Normal CT images. The collection of the data set took 3-4 months. The COVID-19 findings in the images in the dataset have different patterns. In addition most of the images in the data set belong to patients from China, Italy and Spain and have been approved by professionals. The distribution of the CT images of patients in the data set in terms of age and gender are given in Figure 1 and Figure 2, respectively. The images in the data set belong to patients between 10-100 age. According to Figure 1, in the dataset, the images of COVID-19 cases are intense between 40-70 age and showed a normal distribution. The images of normal cases are intense between 50-80 age and showed normal distribution. According to Figure 2, in the dataset, it can be observed that the number of male patients with COVID-19 is slightly higher than that of female patients.

**Figure 1.**
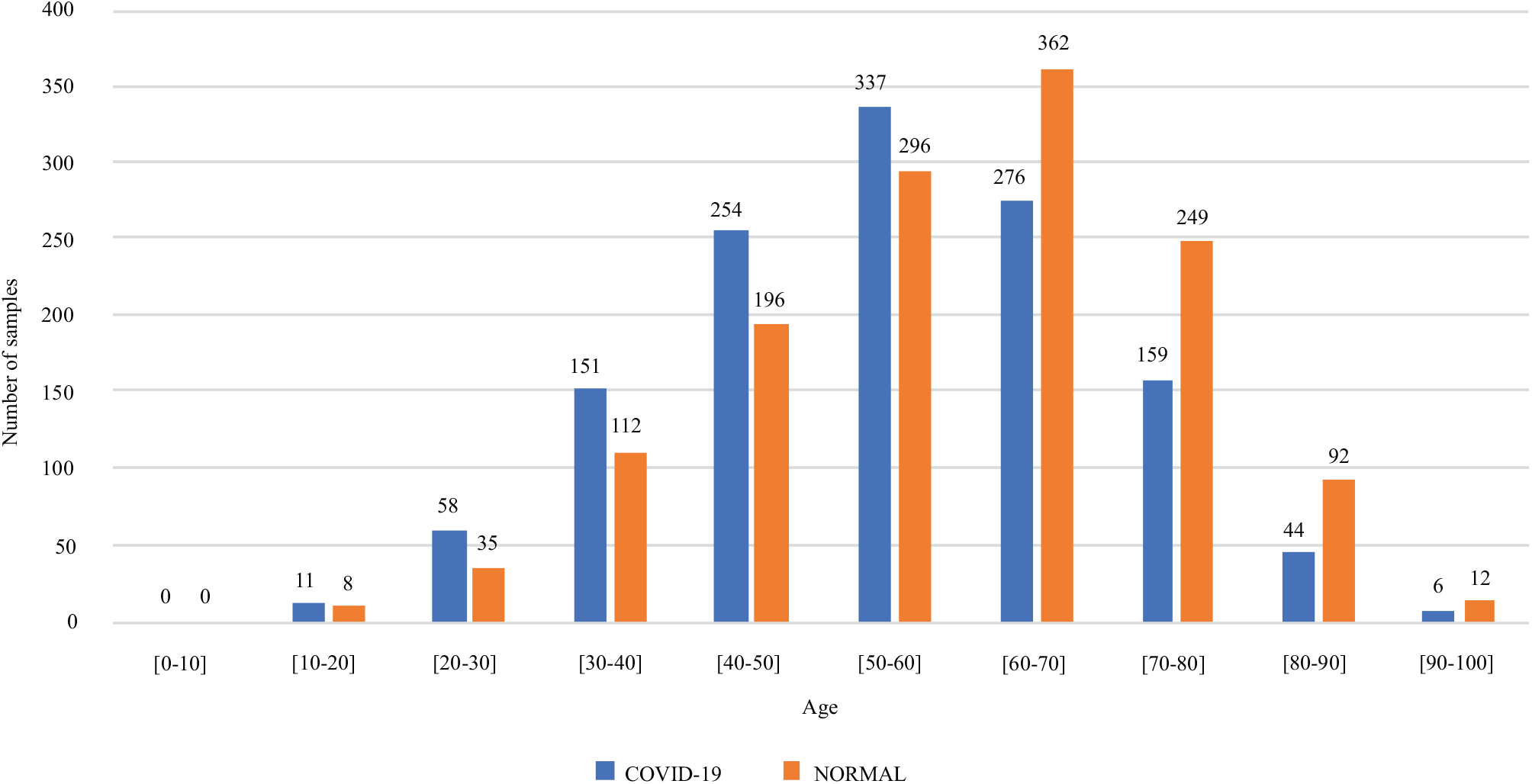
Data distribution in terms of age.

**Figure 2.**
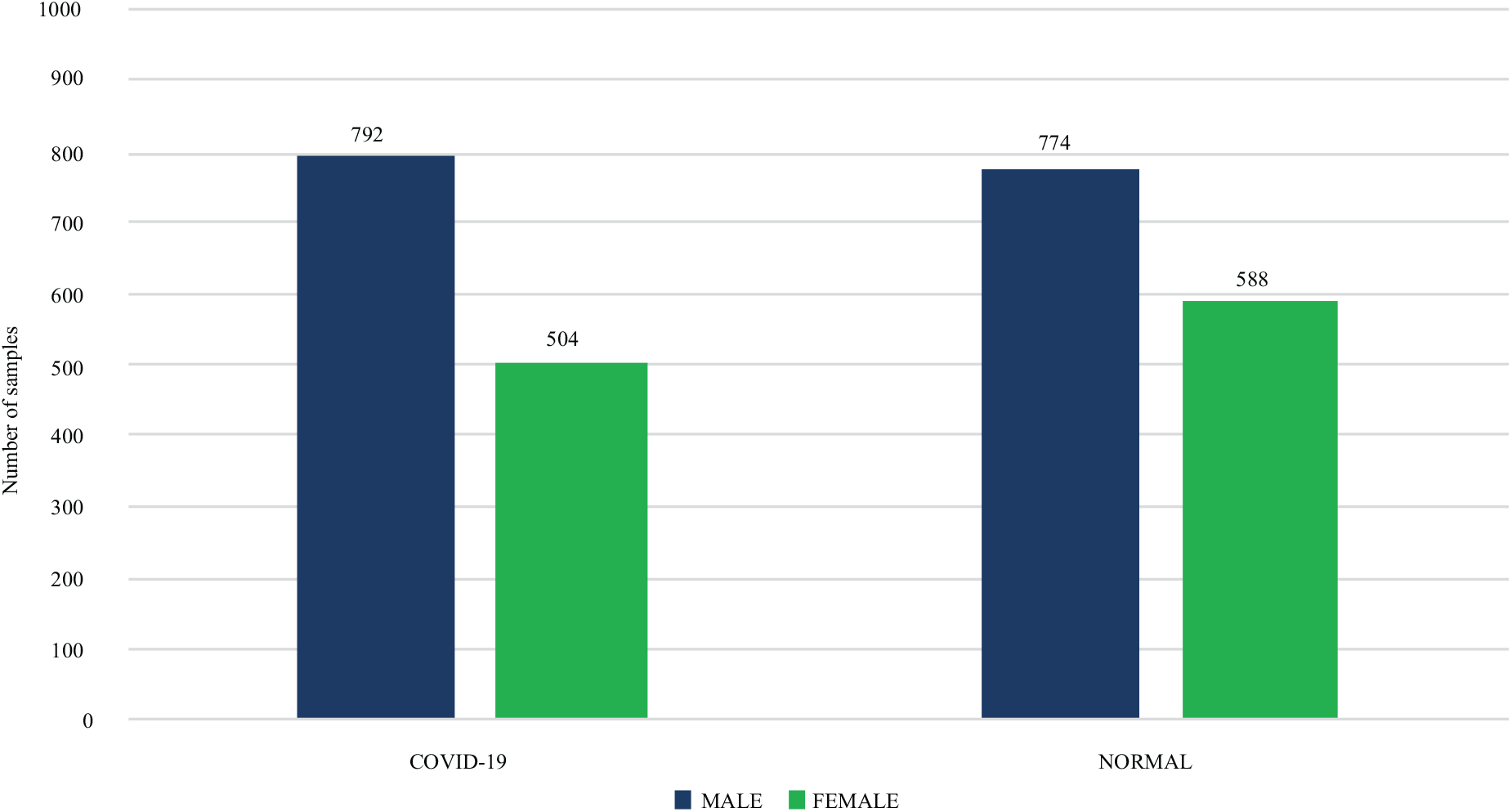
Data distribution in terms of gender.

### 2.2. Simulators and Quantum Computers

The quantum computing simulators we use in our study: Pennylane default simulator^1^ that a noiseless simulator, Qiskit-Aer simulator^2^ that we can define with arbitrary noise rates and noisy Cirq-Mixed simulator^3^ defined with 4 qubit cluster configurations. In addition, near-term 5-qubit IBM Quantum Computers^4^ with different qubit connections and noise rates that we use in our study: IBMQx2 (see Figure 3), IBMQ-London(see Figure. 4) and IBMQ-Rome(see Figure 5).

**Figure 3.**
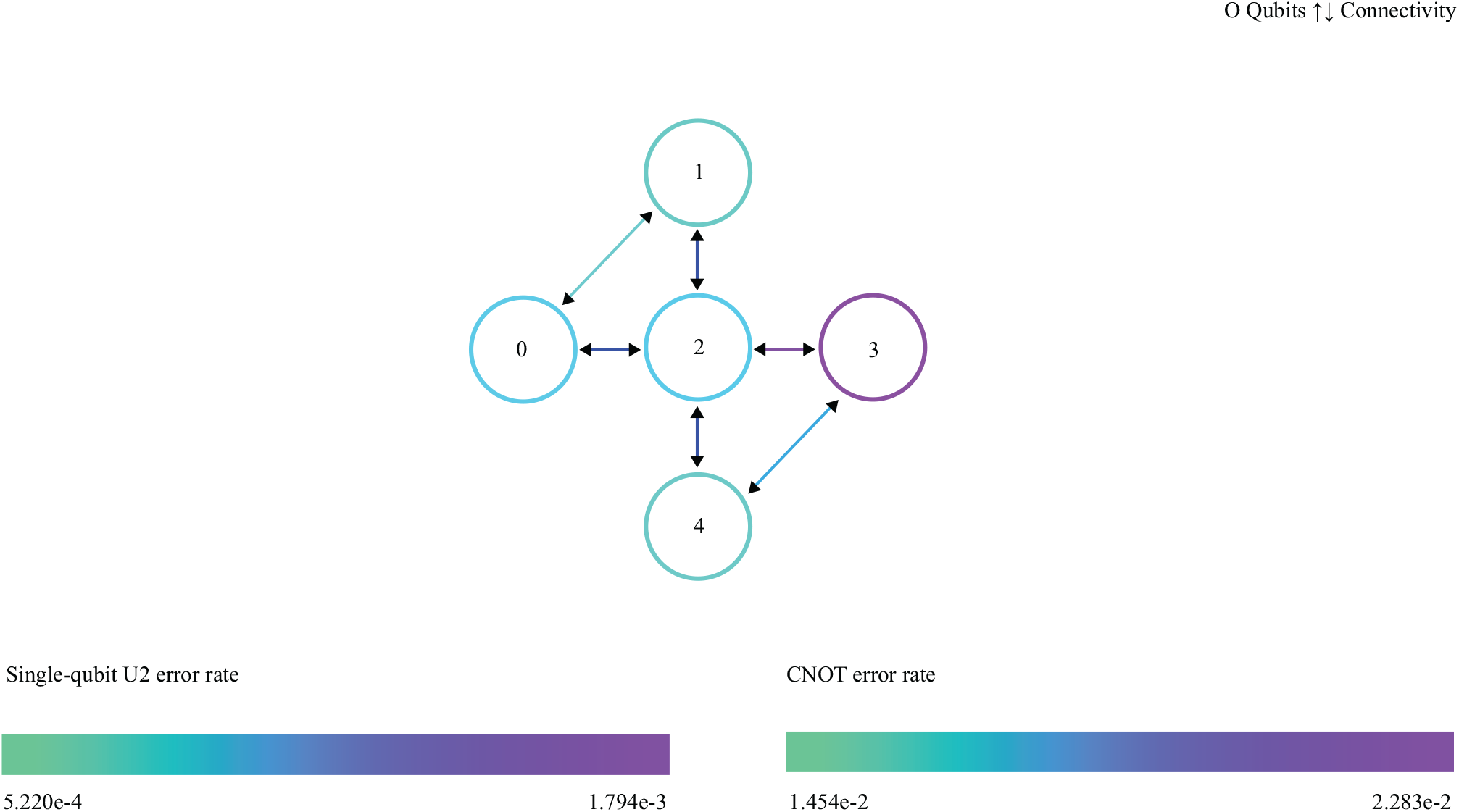
IBMQx2 quantum computer congfigurations.

**Figure 4.**
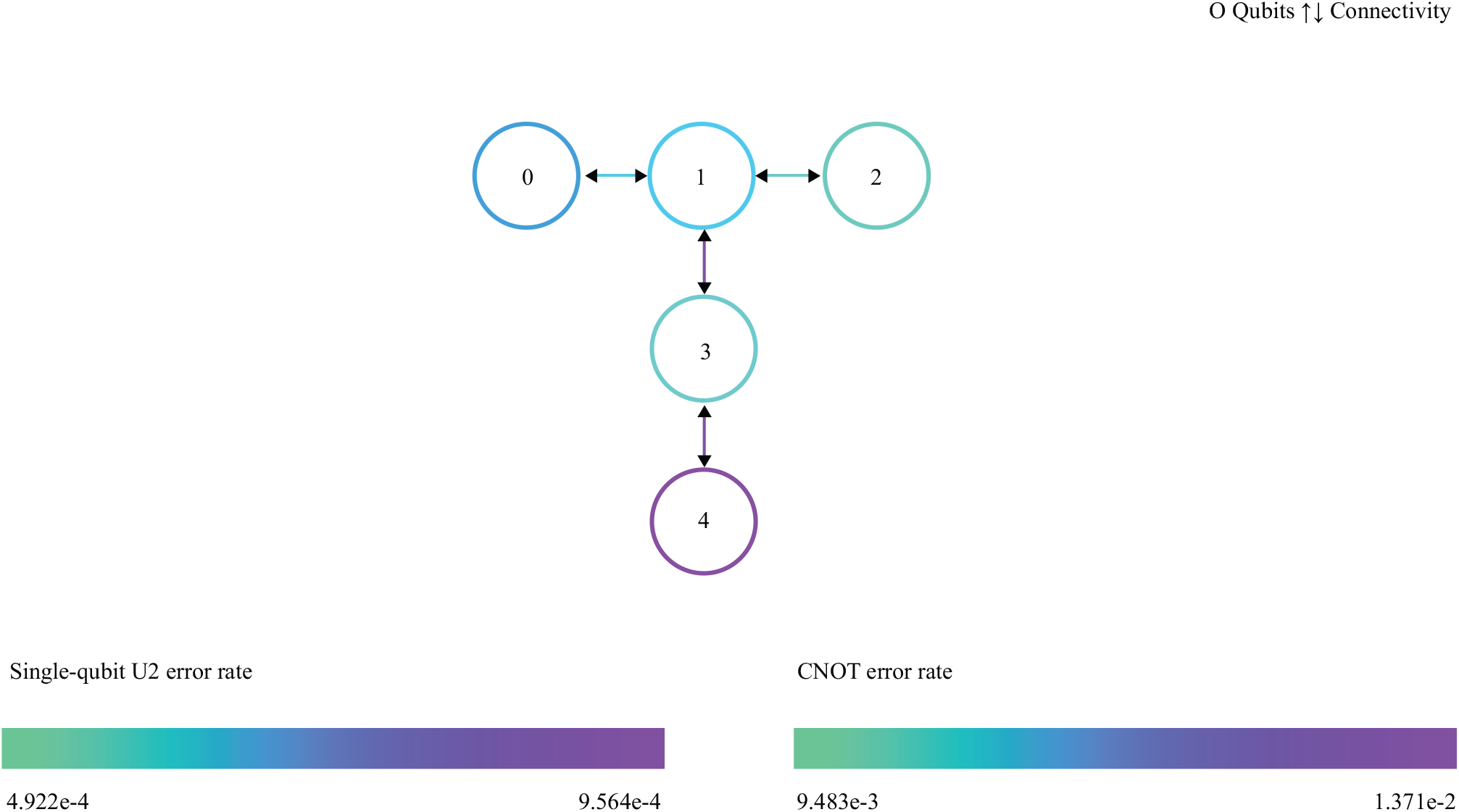
IBMQ-London quantum computer configurations.

**Figure 5.**
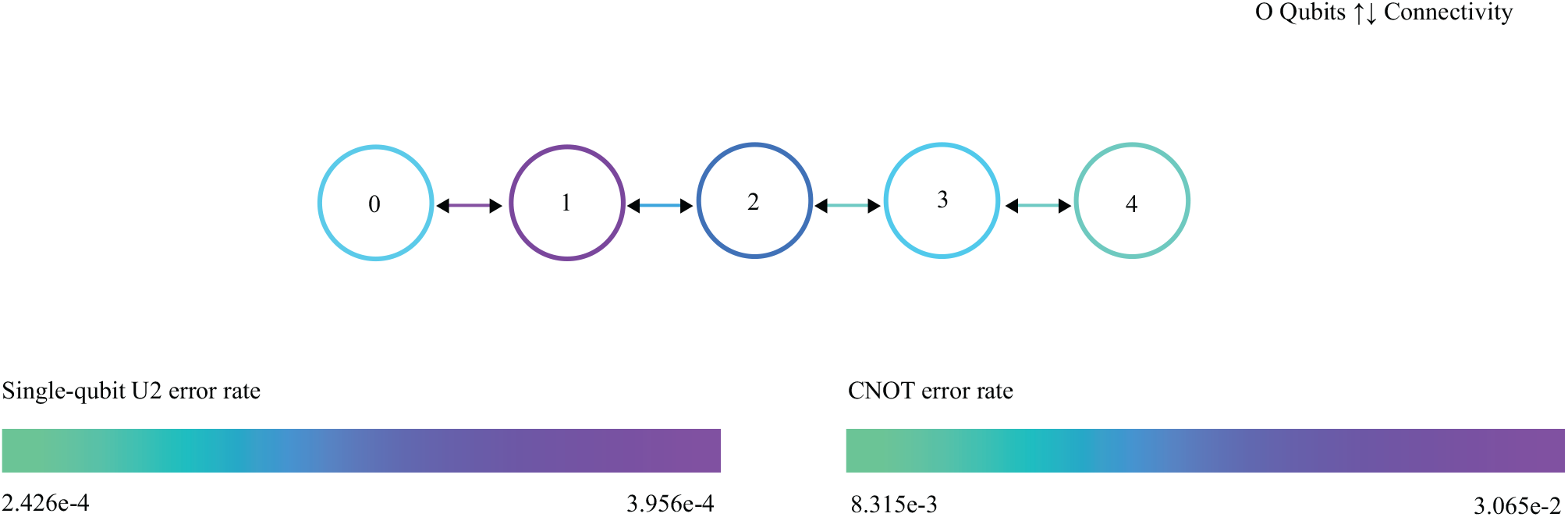
IBMQ-Rome quantum computer configurations.

### 2.3. Preprocessing

Since our data is classic, the preprocessing phase consists of classical operations. The preprocess stage consists of 4 steps. These are given below.

1. Images in the dataset differ in resolution. So we resized it to 3-channel images with 224 × 224 pixels.
2. COVID-19 findings are seen in the lung section of CT images. The information we need is in this section. Therefore, the remaining sections are meaningless and should be removed from the image. We should perform lung segmentation to evaluate the region of interest. We used a method similar to the BConvLSTMU-Net architecture[30] for lung segmentation. We used 1606 CT images and masks for the training of this segmentation network. We initially received the 1e-4 learning rate and dynamically reduced it. We trained using the Adam stochastic optimization algorithm which was developed as a solution to the vanish gradient problem. We obtained the accuracy rate: 0.9902 and the error rate: 0.0325. The ConvLSTMU-Net architecture and example image of output masks is given at Figure 6 and Figure 7, respectively.
3. The region related to the Graphcut[31] image processing method is extracted from CT images. Examples for the images obtained after the Graphcut process are given in Figure 8.
4. Images are normalized according to the mean[0.485, 0.456, 0.406] and standard deviation[0.229, 0.224, 0.225] of the ImageNet data set[32] containing close to 14 million images.

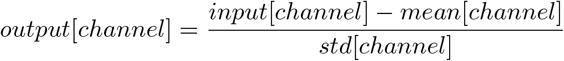

Where *input*[*channel*] is the channel value of the image of the input data. *mean*[*channel*] and *std*[*channel*] is the average and standard deviation channel value of the ImageNet data set.

**Figure 6.**
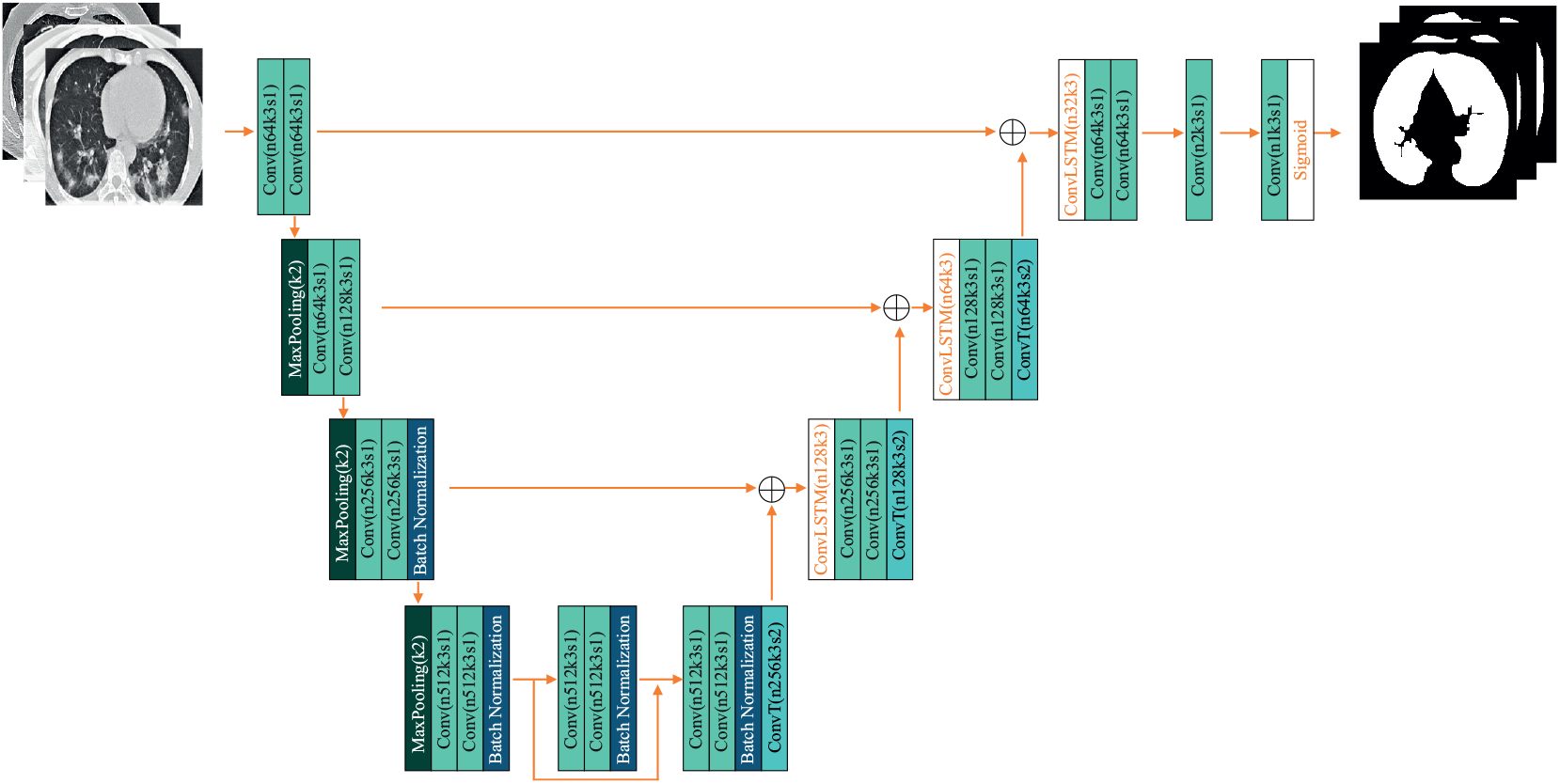
ConvLSTMU-Net architecture.

**Figure 7.**
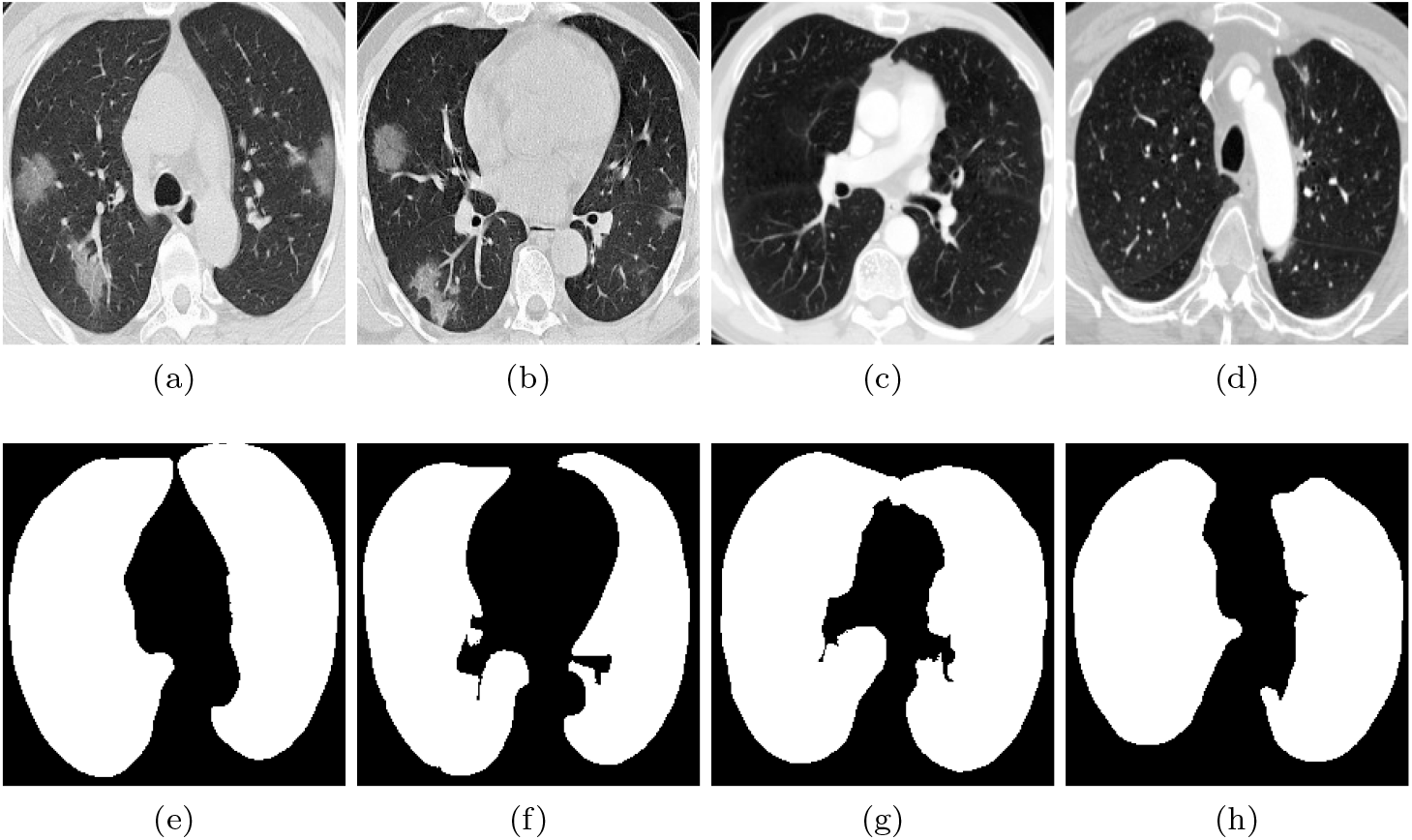
The samples and obtained mask images by using ConvLSTMU-Net model. **a, b, c** and **d** are sample images. **e, f, g** and **h** are mask images.

**Figure 8.**
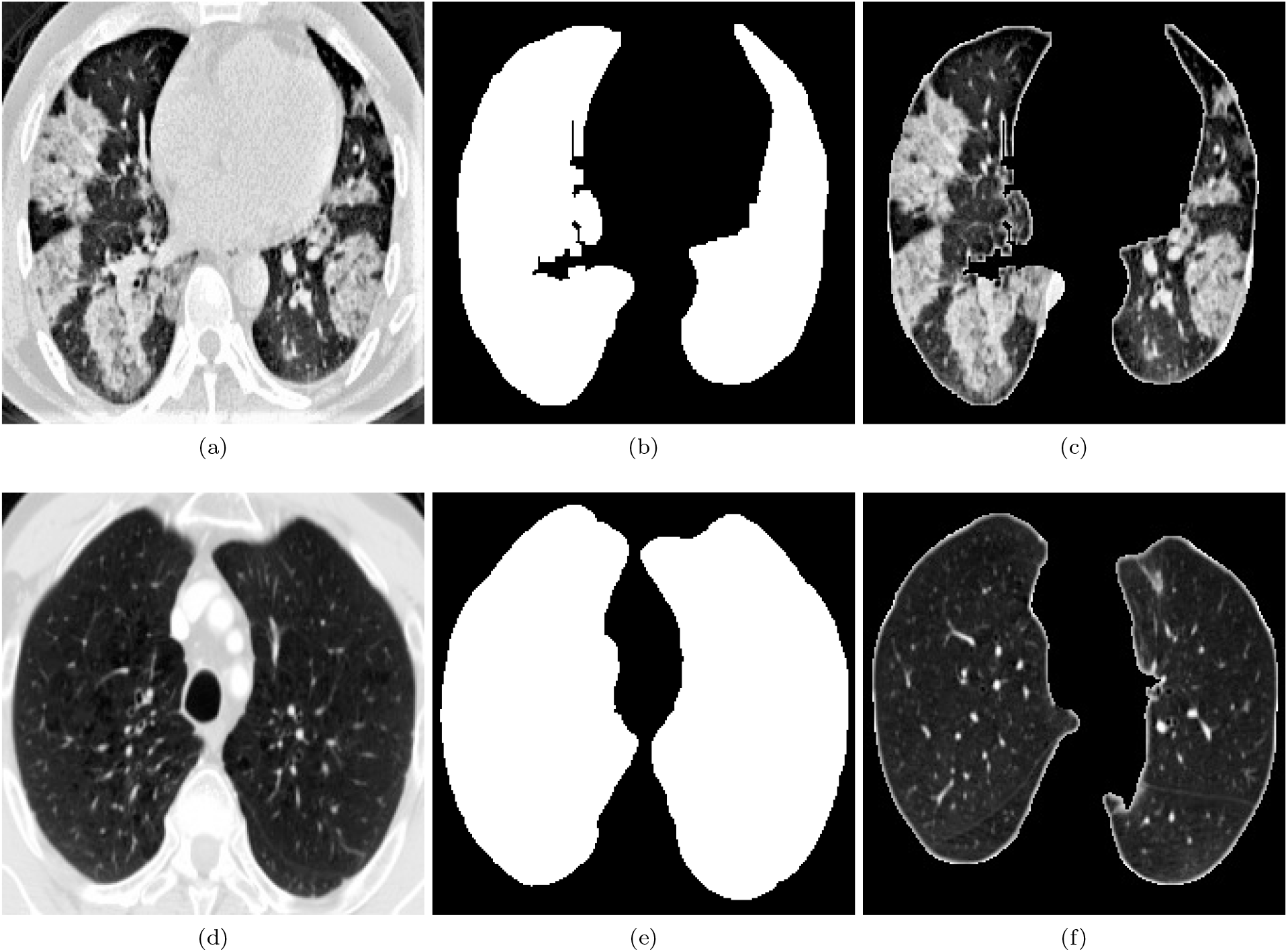
The sample images after applying graph-cut image processing. **a, d** are Sample images. **b, e** are mask images. **c, f** are region of interest images.

### 2.4. Transfer Learning

Transfer learning is an important technique for using artificial neural networks to solve a problem with a small amount of data[6]. If a previously trained artificial neural network is successful in solving a particular problem, it can be reused with some additional training to solve a different but similar problem. Let’s consider a pre-trained deep neural network with the data set used for the solution of a problem. The fully connected layer of the deep neural network is removed from the network. The resulting network can be used as a feature extractor. The end of the pre-trained network that can be used as a feature extractor is added a new network. The new network added by keeping the weights of the feature extractor network fixed is retrained to solve a different or similar problem with a new data set.

### 2.5. Quantum Transfer Learning

Quantum transfer learning is a hybrid machine learning method consisting of a feature extractor classical network and a quantum variational classifier circuit[25]. In our study, we used the ResNet18[33] convolution network as feature extractor. ResNet18 reduces any image to a 512-dimensional feature vector. To classify these features with a 4-qubit variational quantum circuit, we reduced the 512-dimensional feature vector to 4 dimensions with a linear transformation. Variational quantum circuit used as classifier is given in Figure 9. The output of the variational quantum circuit is a 4-dimensional vector. In line with our problem, we reduced the 4-dimensional vector to 2 dimensions. The resulting hybrid model is trained with a 4-qubit variational quantum circuit, by keeping the ResNet18 feature extractor constant with a data set containing COVID-19 positive and COVID-19 negative CT images.

**Figure 9.**
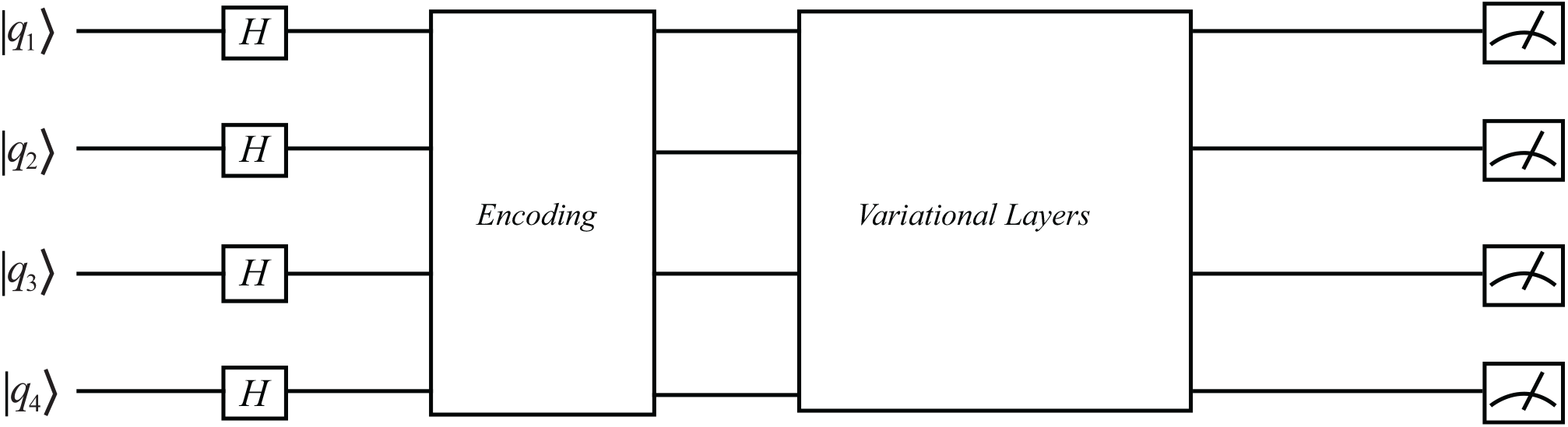
Variational quantum classifier circuit..

### 2.6. Variational Quantum Circuit

The quantum circuit we use for classification consists of 4 stages. The Vartiational quantum circuit[34] is given in Figure 9.

1. All qubits are started in |0) state. Then, Hadamard (*H*) gate is applied to all qubits. Thus, all qubits are brought into superposition.
2. We encoded the classical data together with the *U* circuit by applying additional transformations. We encode the 4-dimensional feature vector with the data encoding circuit consisting of *R*_*y*_(*f*_*i*_) gates and *U* (*α, β, θ, γ*) circuit. The data coding circuit encodes 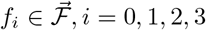 values into the quantum circuit. Where 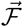 4-dimensional feature vector and *f*_*i*_ is a component of 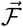 vector. Data coding circuit and U circuit are given in Figure 10 and Figure 11, respectively. In addition, the parameters in the *U* (*α, β, θ, γ*) circuit are trainable.
3. We use a trainable sequence of variational layers consists of an entanglement layer and a data encoding circuit. The entanglement layer is given in Figure 12. The entanglement layer consists of 3 CNOT gates. Makes all qubits entangled. The data encoding circuit used in the variational layer encodes 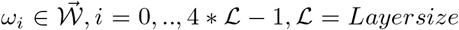 weights. Where 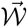is the 4* *ℒ* dimensional weight vector. In our study, we used 6 variational layers by starting the weights randomly.
4. Finally, the measurement process is performed to get the expected value of the 4-qubit relative to the Z operator.

### 2.7. Experimental Evaluation

For the training data set, we use a small number of data with the CT image with 126 COVID-19 tags and 100 Normal(healthy) tags as the opposite of the usual. We trained both a classic model using classical transfer learning and a quantum model using quantum transfer learning using the same training data set. The learning rate is 0.0002 and we used Adam optimization algorithm[35] and Cross Entropy function as activation function. We tested both models with 1170 COVID-19 and 1262 Normal CT images. In addition, we trained the variational quantum classifier circuit in Pennylane, Qiskit and Cirq quantum computing simulators and using IBMQx2, IBMQ-London and IBMQ-Rome quantum computers. We trained the classic model without the quantum variational classifier circuit.

**Figure 10.**
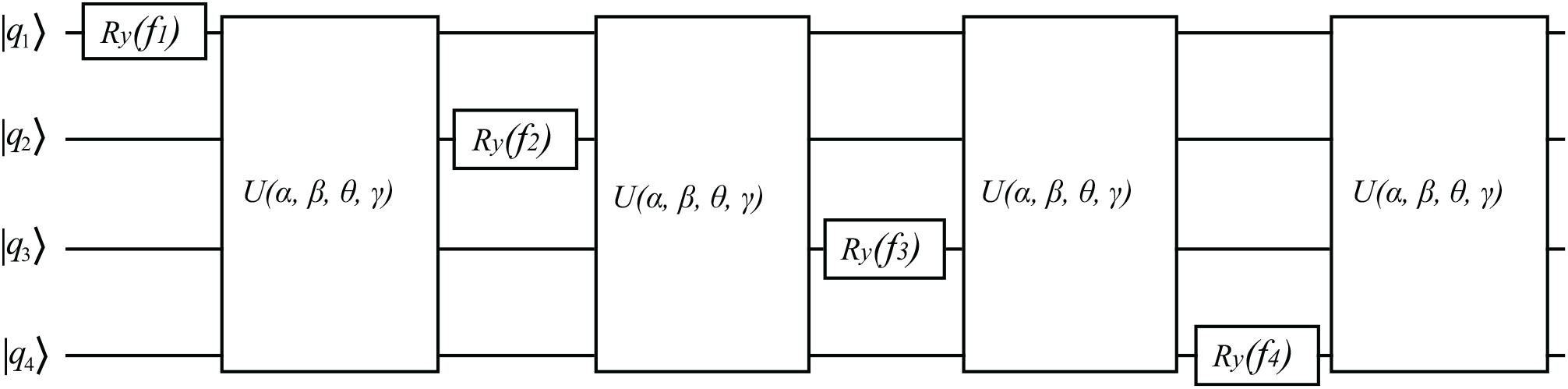
Data encoding circuit.

**Figure 11.**
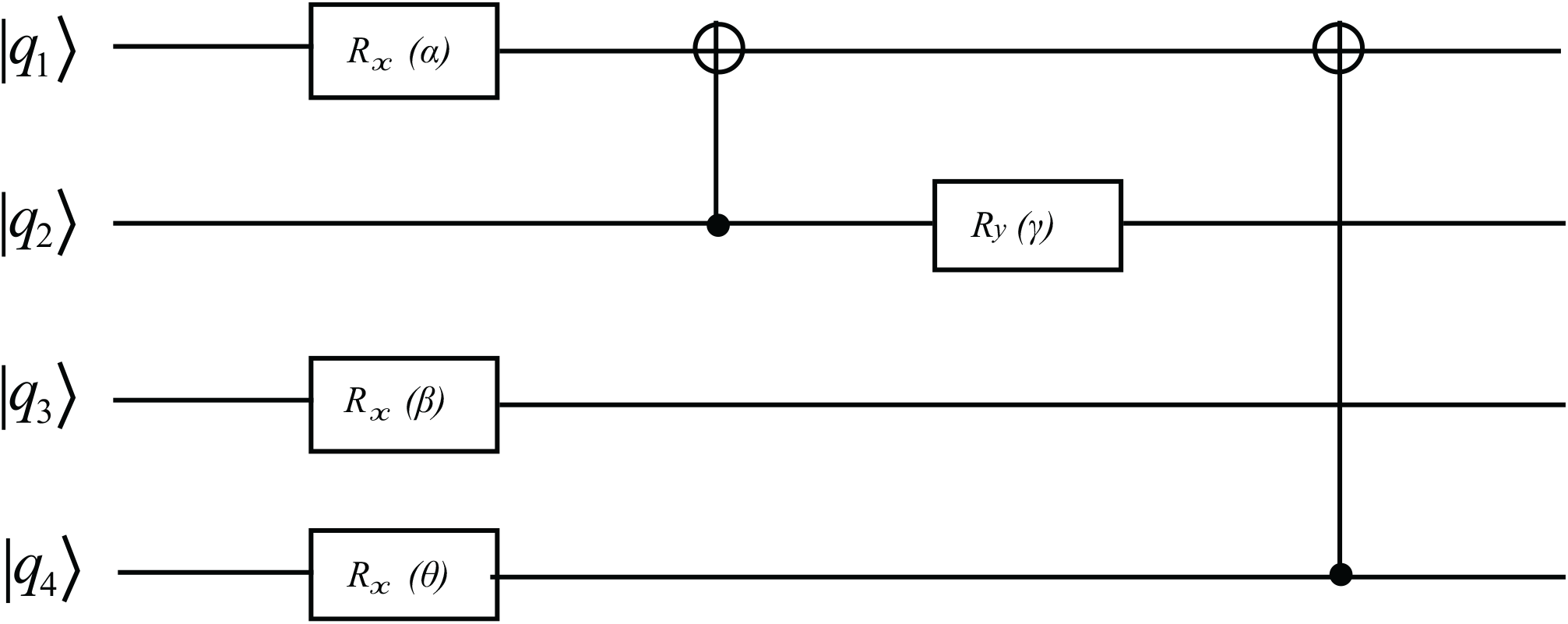
*U* (*α, β, θ, γ*) circuit.

**Figure 12.**
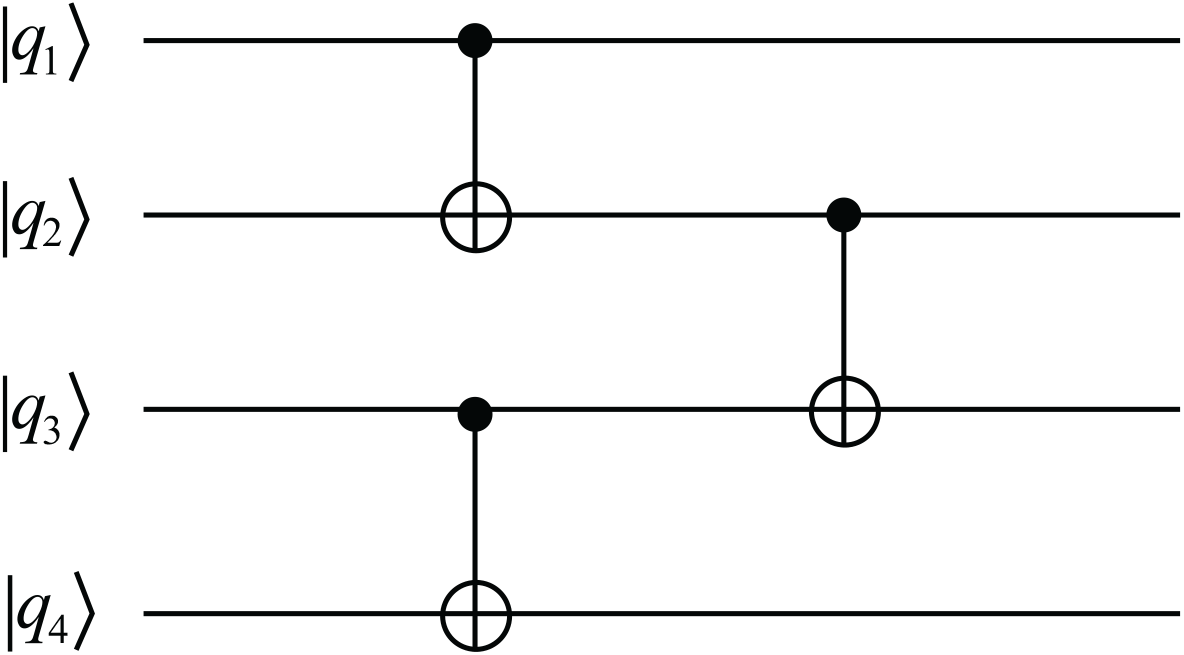
Entanglement layer circuit.

## 3. Results

Performances of the models in this study are measured with metrics such as accuracy(see. Eq 1), precision(see. Eq 2), recall(see. Eq 3), f1-score(see. Eq 4) and specificity (see. Eq 5)[36].

*T*_*P*_ is defined as the number of positive samples estimated correctly, *F*_*P*_ is the number of false predicted samples, *T*_*N*_ correctly estimated number of negative samples, *F*_*N*_ is the number of false predicted negative samples. In this context, accuracy, precision, recall and f1-score are defined as follows. *T*_*P*_ is defined as the number of positive samples estimated correctly, *F*_*P*_ is the number of false predicted samples, *T*_*N*_ correctly estimated number of negative samples, *F*_*N*_ is the number of false predicted negative samples. In this context, accuracy, precision, recall, f1-score and specificity are defined as follows.

- The accuracy rate is an indication of how many of all test data correctly classified and calculated as follows.

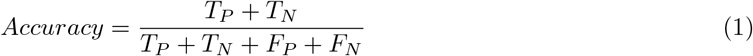
- Precision is the ratio of how many of the COVID-19 cases correctly predicted and calculated as follows.

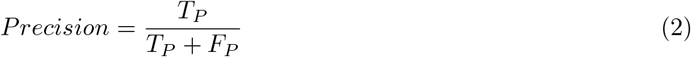
- The percentage of correctly classified labels in truly positive patients is defined as the recall and is calculated as follows.

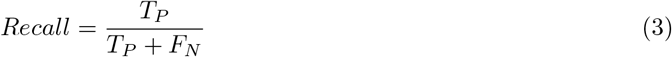
- Recall and precision are two important metrics, and there is a trade-off between them. F1 Score is a good choice when you want to deal with this trade-off and seek a balance between them.

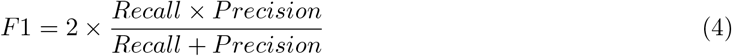
- Specificity is defined as the proportion of true negatives (*T*_*N*_), which got predicted as the negative (or true negative). In other words specificity is how many Normal(healhty) cases are correctly predicted and calculated as follows.

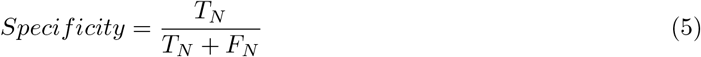

Performance results and confusion matrix of classical model and hybrid quantum model are given in Table 1 and Figure 13 respectively.

**Table 1.** The results show performance results with 95% confidence interval for classic models and hybrid models run on different simulators and IBM quantum computers..

| Models | Accuracy% | Precision% | Recall% | F1-Score% | Specificity% |
| --- | --- | --- | --- | --- | --- |
| Classical Model | $0.909 \pm 0.011$ | $0.926 \pm 0.010$ | $0.890 \pm 0.012$ | $0.908 \pm 0.011$ | $0.929 \pm 0.010$ |
| PennyLane without U | $0.909 \pm 0.011$ | $0.926 \pm 0.010$ | $0.890 \pm 0.012$ | $0.908 \pm 0.011$ | $0.929 \pm 0.010$ |
| **PennyLane with U** | $1.000 \pm 0.000$ | $1.000 \pm 0.000$ | $1.000 \pm 0.000$ | $1.000 \pm 0.000$ | $1.000 \pm 0.000$ |
| Qiskit-Noise Simulator | $0.977 \pm 0.006$ | $0.969 \pm 0.007$ | $0.984 \pm 0.005$ | $0.976 \pm 0.006$ | $0.971 \pm 0.006$ |
| Cirq-Mixed Simulator | $0.956 \pm 0.008$ | $0.970 \pm 0.006$ | $0.940 \pm 0.009$ | $0.951 \pm 0.008$ | $0.971 \pm 0.006$ |
| IBMQx2 | $0.959 \pm 0.008$ | $0.971 \pm 0.006$ | $0.946 \pm 0.009$ | $0.958 \pm 0.008$ | $0.972 \pm 0.006$ |
| IBMQ-London | $0.966 \pm 0.007$ | $0.971 \pm 0.006$ | $0.959 \pm 0.008$ | $0.965 \pm 0.007$ | $0.972 \pm 0.006$ |
| IBMQ-Rome | $0.969 \pm 0.007$ | $0.974 \pm 0.006$ | $0.963 \pm 0.007$ | $0.969 \pm 0.007$ | $0.976 \pm 0.006$ |

**Figure 13.**
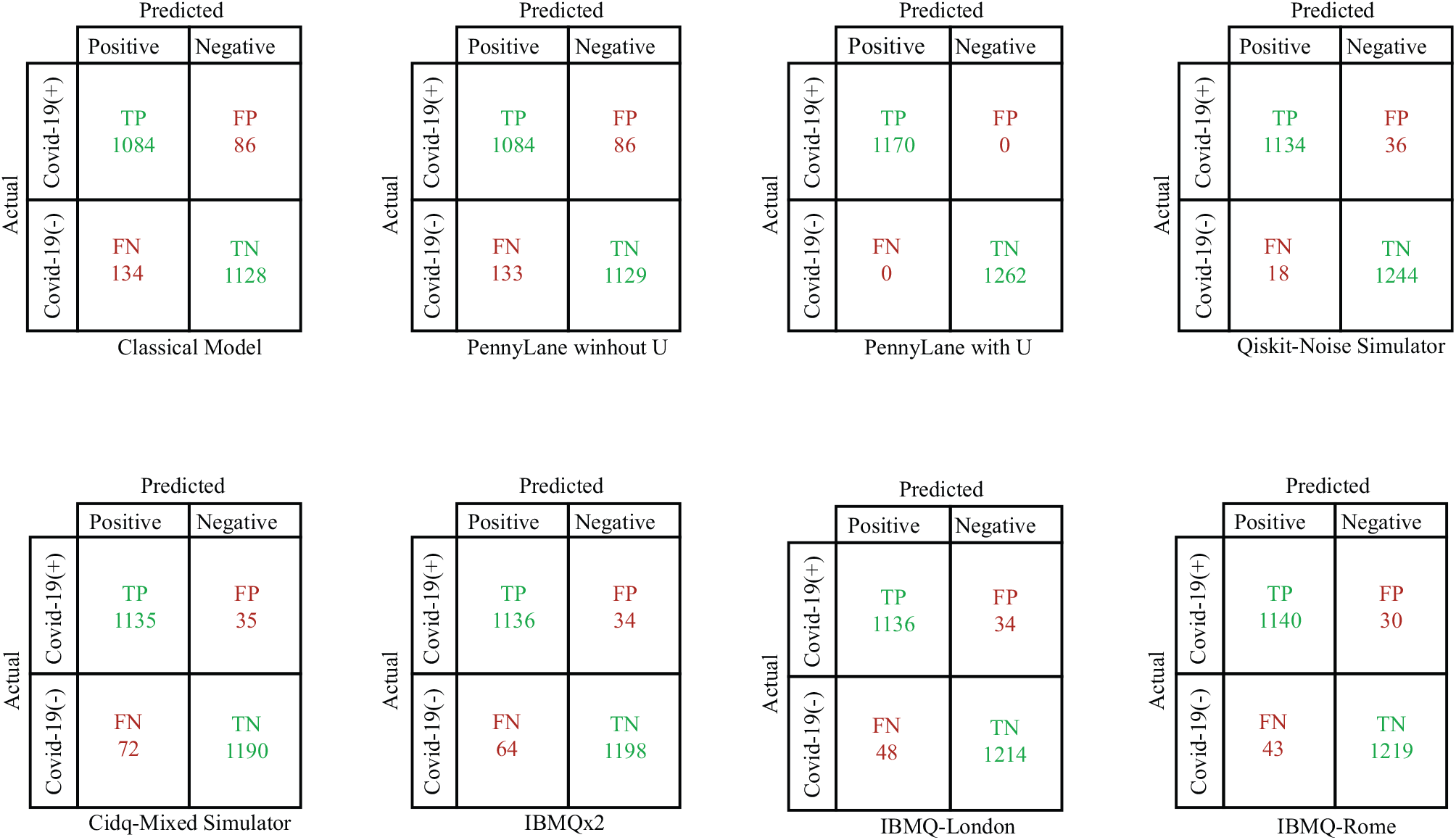
Confusion matrix values of classical model and hybrid quantum model.

## 4. Discussion

Since feature engineering is sensitive and subjective to human bias, an effective feature vector may not be obtained by manually feature extract. Instead, an efficient feature vector is obtained with Convolutional Neural Network(CNN) because CNN-based applications automatically extract properties such as differences, density, color, and shape, and a significant increase in performance has been observed in most studies. The number of data is important for CNNs to be effective, but this problem has been avoided with transfer learning. In our study, a 512-dimensional feature vector is obtained using a pre-trained network, ResNet18. Then the feature vector is encoded into the variational quantum circuit.

By putting all the qubits into the superposition state, all possible states of each component of the feature vector are evaluated. After applying CNOT gates to the qubits in superposition states, all qubits become entangled with each other[12]. In classical computers, while each component of the feature vector is calculated independently from each other, thanks to the quantum computer entangled, each component of the feature vector interacts with each other instantly. The operator affecting one component instantly acts on other components. Thus, different patterns are easily found. Because of all these effects, superposition and entanglement have a direct effect on speed and performance. Thanks to superposition and entangled quantum superior properties, a model can be quickly trained and observed an effective increase in model performance without the need for very large size feature vectors and too many parameter calculations[11, 14, 18, 20–23].

Multiple methods are available to encode classical data into the quantum circuit. Usually, classical data is encoded into base states or amplitudes of quantum states[37]. Another very common method is to embed the classical data into the angles of the Ry rotational gate and encode the amplitudes of the quantum state in the projections of the state occurring on the Bloch sphere on the x and z axes. The resulting situation is classical amplitude. In a way, the third dimension (y-axis) of the Bloch sphere is ignored and treated as if it were a two-dimensional plane. The y-axis of the Bloch sphere contains the imaginary value of the Hilbert space, the complex vector space[38]. This virtual value ensures that quantum states are complex amplitudes. Thus, it is studied in a wider representation space and allows complex entangled situations to be obtained. As a solution to this problem, we encoded the classical data together with the U circuit, applying additional transformations. We also think there is a solution to the problem of over-fitting. On the other hand, we observe an average increase of 5-9% according to performance measurements.

Generally, 70-80% of the data set is reserved for training of machine learning algorithms. It is known that the effect of an increase in education dataset on performance is proportional to classical[39]. On the contrary, we use 20% of the data set for education and 80% for testing.Compared to the conventional model, in the quantum model, we observe there is an average of increase that when using 10% on the noiseless simulator, 8% on noisy simulators and 6% on real quantum computers. When working in small datasets, we can say that quantum is advantageous in terms of performance compared to classical models.

It took about 12-15 minutes to prepare and test the 2432 CT images allocated for testing with a device with a 48GB Ram and Intel Xeon 3.70 GHz processor. With no additional cost or training process for experts, quantum computers are always accessible with the API provided by IBM required^1^.

## 5. Conclusion

It is known that machine learning process, which requires very large computing resources in classical computers and can be done in a long time, can be done in quantum computers in a very short time and with fewer quantum resources. Although it takes a long time because the processes in the quantum processors offered by IBM on the cloud are queued, it is clear that the quantum machines have superiority in terms of performance and speed when compared to conventional computers.

According to the results obtained, it is seen that machine learning processes that require more processing and time in terms of resources and time are performed in a very short time by using only 4-qubit in quantum computers. As can be seen from Table 1, although the size of the data set is small, considering the COVID-19 and Normal classification results on the quantum simulator and quantum real processors according to accuracy, precision, recall, f1-score and specificity (for quantum simulators respectively 95%-100%, 96%-100%, 94%-100%, 95%-100%, 93%-100% and for quantum real processors respectively 95%-96%, 97%, 94%-96%, 95%-96%, 97%) it is seen to be superior to the results on the classic computer (90%, 93%, 89%, 91%, 93% respectively). In addition, we can say by looking at the results in Table 1 that the *U* circuit that we use when encoding the classical data causes the performance results to increase.

Due to the superposition and entanglement, which are the superior features of quantum, we can conclude that quantum computers are advantageous in small size datasets and well classified according to their machine learning performance. If the data are obtained as quantum and the noise rates in the quantum processors are reduced to a minimum, it is clear that these results will be superior both in terms of accuracy and time.

## Data Availability

The data used in the study were collected from open source data sets.

## Acknowledgment

This work was produced within the framework of Erdi Acar’s master thesis. We would like to thank anonymous referees for valuable suggestions.

1 RADIOPEDIA(2005). [online]. Website www.radiopaedia.org [accessed 01 04 2020]

2 SIRM(2019). COVID-19 Database [online]. Website https://www.sirm.org/en/category/articles/covid-19-database [accessed 01 04 2020]

3 GITHUB(2020). COVID-CT [online]. Website www.github.com/UCSD-AI4H/COVID-CT [accessed 01 04 2020]

4 MEDSEG(2019). COVID-19 CT segmentation dataset [online]. Website www.medicalsegmentation.com/covid19 [accessed 15 04 2020]

5 RADASS(2018).[online]. Website www.radiologyassistant.nl [accessed 10 04 2020]

6 KAGGLE(2010). COVID19-CT-Scans [online]. Website www.kaggle.com/andrewmvd/covid19-ct-scans [accessed 20 04 2020]

7 KAGGLE(2010). COVIDCT [online]. Website www.kaggle.com/luisblanche/covidct [accessed 20 04 2020]

1 PENNYLANE(2019). Pennylane Simulator [online]. Website www.pennylane.ai [accessed 15 05 2020]

2 QISKIT(2019). Qiskit Simulator [online]. Website www.qiskit.org [accessed 15 05 2020]

3 CIRQ(2019). CIRQ Noisy Intermediate-Scale Quantum Simulator [online]. Website www.cirq.readthedocs.io [accessed 15 05 2020]

4 IBMQ(2019). IBM Quantum Computers [online]. Website www.quantum-computing.ibm.com [accessed 15 05 2020]

1 IBMQ(2019). IBM Quantum Computers [online]. Website www.quantum-computing.ibm.com [accessed 15 05 2020]

